# Identification of a novel proteomic Biomarker in Parkinson’s Disease: Discovery and Replication in Blood, brain and CSF

**DOI:** 10.1101/2021.12.26.21268282

**Authors:** Laura Winchester, Michael Lawton, Imelda Barber, Jessica Ash, Benjamine Liu, Samuel Evetts, Lucinda Hopkins-Jones, Suppalak Lewis, Catherine Bresner, Siv Vingill, Ana Belen Malpartida, Nigel Williams, Steve Gentlemen, Richard Wade-Martins, Brent Ryan, Alejo Holgado-Nevado, Michele Hu, Yoav Ben-Shlomo, Donald Grosset, Simon Lovestone

**Author notes:** Corresponding Author: Laura Winchester. Currently at Otsuka Pharmaceuticals (Jessica Ash). Currently at TrialSpark (Benjamine Liu). Currently at Janssen (Simon Lovestone).

## Abstract

Biomarkers to aid diagnosis and delineate progression of Parkinson’s Disease (PD) are vital for targeting treatment in the early phases of disease. Here, we aim to discover a multi-protein panel representative of PD and make mechanistic inferences from protein expression profiles within the broader objective of finding novel biomarkers.

We used aptamer-based technology (SomaLogic^®^) to measure proteins in 1,599 serum samples, 85 CSF samples and 37 brain tissue samples collected from two observational longitudinal cohorts (Oxford Parkinson’s Disease Centre and Tracking Parkinson’s) and the PD Brain Bank, respectively. Random forest machine learning was performed to discover new proteins related to disease status and generate multi-protein expression signatures with potential novel biomarkers. Differential regulation analysis and pathway analysis was performed to identify functional and mechanistic disease associations.

The most consistent diagnostic classifier signature was tested across modalities (CSF AUC = 0.74, p-value = 0.0009; brain AUC = 0.75, p-value = 0.006; serum AUC = 0.66, p-value = 0.0002). In the validation dataset we showed that the same classifiers were significantly related to disease status (p-values < 0.001). Differential expression analysis and Weighted Gene Correlation Network Analysis (WGCNA) highlighted key proteins and pathways with known relationships to PD. Proteins from the complement and coagulation cascades suggest a disease relationship to immune response.

The combined analytical approaches in a relatively large number of samples, across tissue types, with replication and validation, provides mechanistic insights into the disease as well as nominating a protein signature classifier that deserves further biomarker evaluation.

## Introduction

Parkinson’s disease (PD) is a complex neurological disorder resulting in disabling motor and non-motor deficits, often accompanied by cognitive impairment, as a consequence of loss of dopaminergic neurons within the substantia nigra pars compacta and ventral tegmental area [1]. Given the prevalence of over 350 per 100,000 people globally which increases with age [2], PD represents a heavy burden not only to patients but to families, carers and society. It has become increasingly apparent that there is a prolonged prodromal phase of PD [3], similar to other neurodegenerative conditions such as Alzheimer’s disease (AD) [4]. A biomarker(s) for use in selection and stratification in clinical trials and early therapeutic intervention has become an increasingly pressing objective for research as we seek disease-modifying therapies. In AD, fluid biomarkers of pathological processes have been identified in both cerebrospinal fluid (CSF) and blood and are increasingly used in clinical trials [5]. In PD, reliable biomarkers for use in clinical settings that recapitulate underlying pathophysiology are rare, making the search for candidate protein markers more urgent [6]. Basso *et al*. [7] described 44 proteins that supported the role of oxidative stress. Subsequently, Chen-Plotkin *et al*. [8] used immunoassays of 151 proteins in plasma and found epidermal growth factor (*EGF*) to be associated with cognitive decline in PD. In addition to targeted biomarker testing, biomarker identification has been broadened to include unsupervised discovery approaches. Chen-Plotkin used the SomaLogic aptamer capture technology to identify a putative PD related signature, finding four new markers of PD which were able to differentiate PD from Amyotrophic Lateral Sclerosis patients [9]. Serum neurofilament light chain levels have been shown to be raised in PD compared to controls, with longitudinal data showing positive associations with motor scores [10]. These results and those from other studies suggest that there might be a signature of disease in peripheral fluids such as blood, with potential as use as a biomarker [6,11].

We sought to build on past studies and learn from similar approaches in other neurodegenerative diseases [12–14] to identify protein-based biomarkers from peripheral fluids in PD in larger cohorts. We used high dimensionality proteomics technology (SOMAmer assay, SomaLogic^®^) based on aptamer capture assays of between 1000 to 4000 proteins. Here, we describe a dual approach to the study analysis; a machine learning approach to discover a reproducible multi-protein signature representative of PD and an in-depth analysis of proteomic function to provide mechanistic disease insights.

## Methods

### Study Design and Participants

Parkinson’s disease cases and control samples for this study were obtained from three cohorts; the Oxford Parkinson’s Disease Centre (OPDC) Discovery Cohort [15], the Tracking Parkinson’s cohort (Parkinson’s Repository of Biosamples and Networked Datasets - PRoBaND) [16] and the Parkinson’s UK Brain Bank (PUKBB) [17]. Samples were organised and tested by the Mapping Proteomics to Parkinson’s disease UK (MAP2PD-UK) collaboration with a multi-stage design including blood, brain and CSF samples (Figure 1). Cohort details were as follows:

**Figure 1.**
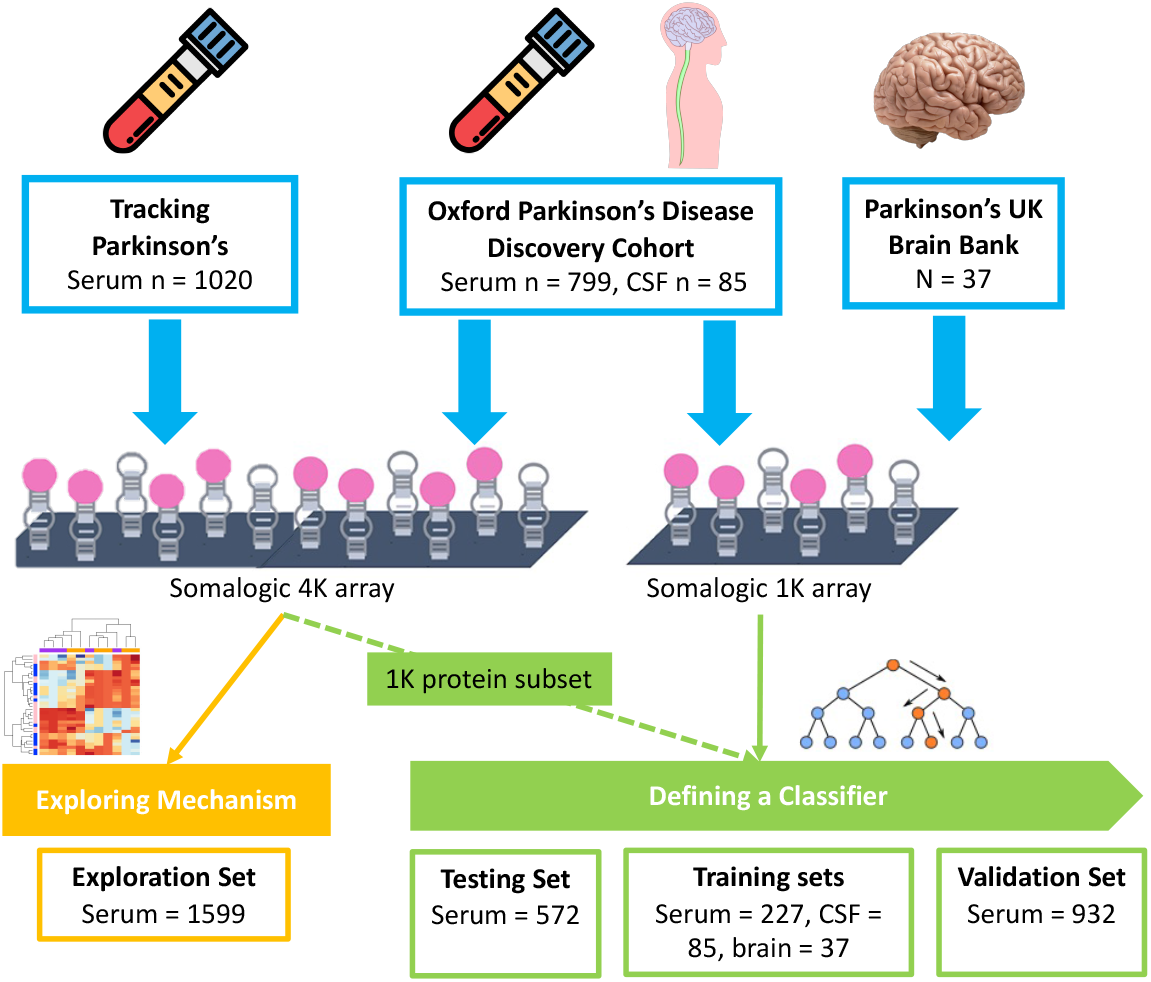
The Mapping Proteomics to Parkinson’s disease UK Project - Multiple Cohort Study Design. Key Brain: Brain tissue, CSF: Cerebrospinal fluid, Serum: Blood derived serum sample

#### Oxford Parkinson’s Disease Centre (OPDC) Cohort

The OPDC Discovery cohort was collected from eleven hospitals covering a UK Thames Valley population of 2.1 M. Cases were selected using clinical data permitting endophenotype analysis by the severity of motor phenotype (both measured using MDS-UPDRS scores) and by cognitive impairment (measured using Montreal Cognitive Assessment, MoCA). Individuals with an alternative diagnosis or with a clinical impression of PD probability < 90% at their latest visit were excluded. Levodopa equivalent daily dose (LEDD) was recorded at the time of sampling. All individuals with a baseline disease duration of > 3.5 years were excluded. For the initial sampling, using the OPDC-Serum-1K, disease free controls were matched by gender and age, where age matching was within ± 2.5 years. Individuals with common LRRK2 or GBA mutations were excluded. The OPDC-Serum-1K set contained 144 samples including 65 controls. CSF proteins were measured for 85 samples. The OPDC-Serum-4K set contained 400 cases and 172 controls, here the larger sample set incorporated all available controls and mutation exclusion filters were not required.

#### Tracking Parkinson’s cohort

The Tracking Parkinson’s (TP) or PRoBaND cohort is a prospective study spanning 72 UK locations. Two sets of samples were selected from this cohort. An initial set (TP-Serum-1K) and a furthermore comprehensive set (TP-Serum-4K). The TP-Serum-1K set contained 80 cases and was combined with the OPDC-serum-1K for analysis. Age matching was within 2.5 years. The TP-Serum-4K sample set (n=1,020) includes a broader range of endophenotypes from the clinical measures. All individuals with a baseline disease duration of > 3.5 years were excluded. Individuals with an alternative diagnosis or with a clinical impression of PD probability <90% at their latest visit were excluded. In both sample sets LEDD was recorded at the time of sampling.

#### Parkinson’s UK Brain Bank (PUKBB)

The Parkinson’s UK Brain Bank (PUKBB) is part of the Multiple Sclerosis Society and Parkinson’s Tissue Bank based at Imperial College London. 37 brain (anterior cingulate cortex) samples were used for the Brain test set, all originating from the PUKBB cohort. 24 brain samples were from PD donors. 13 brain samples were from control donors. Samples were selected which had a post-mortem interval of < 26 hours and RNA integrity of > 6.0.

### Classifying Disease Severity in the Cohorts

Case samples were split into ‘mild’ and ‘severe’ phenotypes, defined by cognitive and motor symptoms in order to make inferences about changes in disease pathology during disease progression. Initially, for OPDC - Serum - 1K subset, samples were selected using the bottom 15% (mild) and the top 10% (severe). For other phases of the analysis an equivalent approach was generated to generate a severity score per patient. The MoCA and MDS UPDRS III scores were standardised by square root transformation (Equation 1 A and B), then combined to generate a severity score per patient. This score was used to sort samples by quantiles: mild, intermediate and severe (quantiles were defined at 15% and 85%). The severity score was used as a continuous measure in the larger validation set.

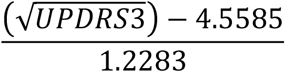

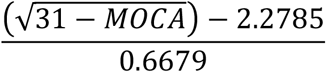

### Protein measurements

All samples analysed in this study had protein levels measured using a Slow Off-rate Modified Aptamer (SOMAmer)–based capture array in five batches using this evolving array (‘SOMAscan’ assay from SomaLogic, Inc. Boulder, CO)[18]. Over the course of the study this platform was upgraded - namely, OPDC and brain samples were run on the Version 2 panel which includes 1,129 proteins, Tracking Parkinson’s and CSF samples were run on an updated panel (Version 3) with a total of 4,006 proteins including those on Version 2. Protein concentration measurements were derived from Relative Fluorescent Units (RFU), which had undergone Somalogic proprietary experimental normalisation to standardize noise between sample plates.

### Quality Control and Data Pre-processing

Samples not meeting quality control standards defined by SomaLogic (acceptance criteria concentration range 0.4-2.5 based on experimental controls) were removed from further analysis. For classifier discovery 1,129 protein assays were used which included multiple isoforms for a single protein in some cases. For differential regulation and enrichment analysis 3,381 assays were used, each protein represented once. The per cohort sample and assay pass rate summary is included in Supplementary Table 1. All data pre-processing and analysis was performed using the statistical coding language R (version 4.0.2). Potential experimental confounders for these cohorts were checked for significant effects (e.g., assay plate, location, submission box, cohort, submission batch year). Principal Component Analysis was used to understand underlying variation, and where confounders were related to principal components a further step was introduced. Visual inspection was used to assess whether any of the recorded experimental variables were associated with any of the first principal components, highlighting cohort and array type as confounders. After this step, two different pre-processing strategies were used depending on the final objective. When the final objective was biomarker discovery protein concentrations were normalised per data subset. When the final objective was differential expression analysis, protein concentrations were normalised using a bulk strategy.

### Targeted Protein Analysis

A narrative review of PD biomarker studies was conducted to select a range of diagnostic and prognostic proteins. This resulted in a list of 50 proteins, although not all were represented in our Somalogic platform. Protein expression was log2 transformed. Finally, a list of 25 unique proteins (30 assays total) were taken forward for univariate analysis using a simple logistic regression (disease status) or linear regression (MoCA and MDS-UPDRS III) on each sample subset. As this method focused on selected proteins to test known biomarkers, p-values are presented as independent results without adjustment.

### Classification of a biomarker

#### Biomarker Method Quality Control

The classification pipeline incorporated all the samples measured during the project, using only those proteins common to versions 2 and 3 of the SomaLogic platform (n=1,047). To allow the unbiased inclusion of all sample type modalities, we limited the selection to the 1,047 proteins found on the version 2 SomaScan array. Sample sets were split by batch and cohort for initial quality checks before assignment into testing and training groups as described in Figure 1. Each dataset was corrected for unwanted variation due to demographics by building a generalised linear model per protein, and then retaining the residuals calculated by the model. In this case, these generalised linear models used the to-be-corrected protein level as outcome, while the predictor variables were the covariates. The covariates were cohort (Serum-1K only), participants’ LEDD at time of sampling, date samples were tested on the SOMAscan assay (OPDC-CSF only) and the date the samples were prepared (Brain only). Following correction, each dataset was z-score standardised.

#### Discovery and Classification

We first performed a two-stage machine learning analysis – discovery and replication to identify a multi-protein signature of disease using a case-control design. In the OPDC-serum dataset (‘discovery’ set) we ranked all proteins using simulated annealing as implemented in R package ‘caret’ (version 6.0.77) [19]. This iterative algorithm starts with a random selection of proteins, then randomly perturbs the selection, and keeps the new list of proteins only if it is better at discriminating cases from controls with a generalised linear model. Random perturbation is repeated iteratively until the algorithm converges into a final list of proteins. Proteins are then ranked according to how often they are selected for their discriminative power. In the following step, using the resulting ‘N’ top ranked proteins, a random forest classifier was built (R package ‘caret’) to discriminate between cases and controls. This was repeated with increasing values of N, from 1 to 200, creating a total of 200 random forest classifiers, and recording in each case the accuracy that the classifier achieved.

For the replication stage, the identified protein classifiers were tested on three independent datasets, each representing three different tissue types, serum (Serum-1K), CSF (OPDC-CSF) and post-mortem brain (Brain). The classifiers output from the replication stage were taken as a refined protein panel set.

Classifiers were compared using the area under the receiver operator characteristic curve (AUC), which was calculated using the function ‘predict’ as implemented in the R base package ‘stats’, and the ‘prediction’ and ‘performance’ functions as implemented in the R package ‘ROCR’ (version 1.0.7). The p-value associated to this AUC was calculated using 10,000 random permutations of disease status to create a distribution of the statistic. Optimising biomarker discovery methodology in a multi-modal cohort is key to finding the set of proteins that most robustly predicts the outcome. In addition to in depth quality control (Supplementary Methods), selection of the correct sample set for training is important. Three sample sets were tested, OPDC serum, TP serum and CSF (OPDC). Our biomarker was required to perform well in all modalities (Supplementary Table 2). The strongest AUC after training were for CSF and brain. However, the priority for this analysis was for a good prediction AUC in serum testing. The serum training set selected was from a single cohort (OPDC) and contained similar case-control proportions to the testing datasets (70 - 30 %).

### Validation of the novel multi-protein signature

The best multiprotein classifier, or signature, was then explored further in the validation data set, which consisted of serum samples from Tracking Parkinson’s. Protein expression data was log^2^ transformed as a standard experimental normalisation strategy. Two approaches were used to convert the signatures from the protein classifier to a single indicator. First, a mean expression per sample with z-score transformation was used. Secondly, singular value decomposition was used to generate an eigengene representation of the signature in an approach to reduce dimensionality. The performance of these classifiers or signatures in CSF (total protein = 142), brain (total protein = 89) and serum (n = 31) were assessed using association to disease status as outcome.

### Protein Differential Regulation and Enrichment Analysis

In order to explore disease mechanisms, we focused on the data generated from the version 3 Somalogic assay, which included 4,006 protein assays. Protein expression values were log^2^ transformed. This analysis used 1,599 serum samples with a matched case-control structure from the two longitudinal cohorts. An additional quality control step was undertaken to reduce between-cohort effects. ComBat normalization [20] was employed to remove this structure, while measuring dataset structure was checked using Uniform Manifold Approximation and Projection (UMAP) clustering. We performed differential regulation analysis on log^2^ transformed data using linear models corrected for age (continuous), gender, and cohort. All generalised linear models were created using the R function ‘glm’ with linkage function binomial for disease status and gaussian for continuous phenotypes (e.g. MoCA and MDS-UPDRS). For pathway enrichment analysis Over Representation Analysis (ORA) was implemented in the R library clusterProfiler [21] using the KEGG and GO databases whereby proteins signatures are tested (hypergeometric) to understand whether a function or pathway is enriched more than would be expected by chance.

Comprehensive co-expression analysis was performed using Weighted Gene Correlation Network Analysis (WGCNA) [22]. The standard pipeline was adapted for use with the proteins as described by Seyfried *et al*. [23]. Briefly, modules were capped at minimum protein number = 30, power at 2 and the block-wise adjustment step was incorporated to reduce module noise and increase the robustness of protein selection. ORA was implemented on each module to understand functionality and relevance to disease mechanism. The eigengene value for each module was compared to clinical measures using logistic regression (PD case status) and general linear model (MoCA and MDS-UPDRS III) to understand the relationship to disease.

## Results

Using an aptamer capture assay measuring over 1000 proteins we sought to find signatures of disease that might function as biomarkers and provide insights into disease mechanisms. To do this we utilised serum samples from two cohorts, the Oxford Parkinson’s Disease Centre Cohort (OPDC) and Tracking Parkinson’s, together with CSF samples from OPDC and brain samples from the Parkinson’s UK Brain Bank (PUKBB). The participant characteristics from these datasets are described in Table 1. The ratio of cases to controls across samples varied from 82% to 65% with cases predominating. As expected, there was a male predominance and the mean age for the UKBB was from 9 to 13 years older.

**Table 1.**
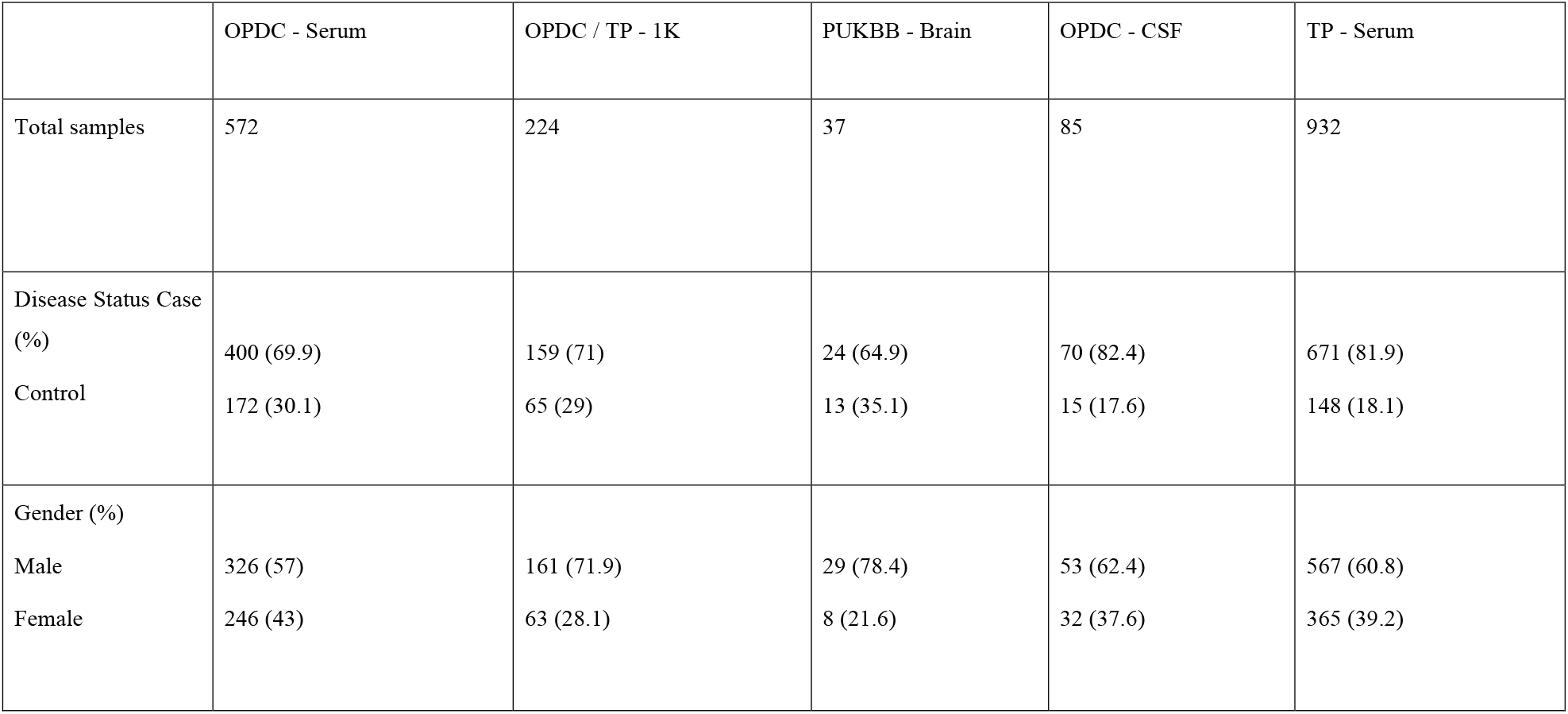
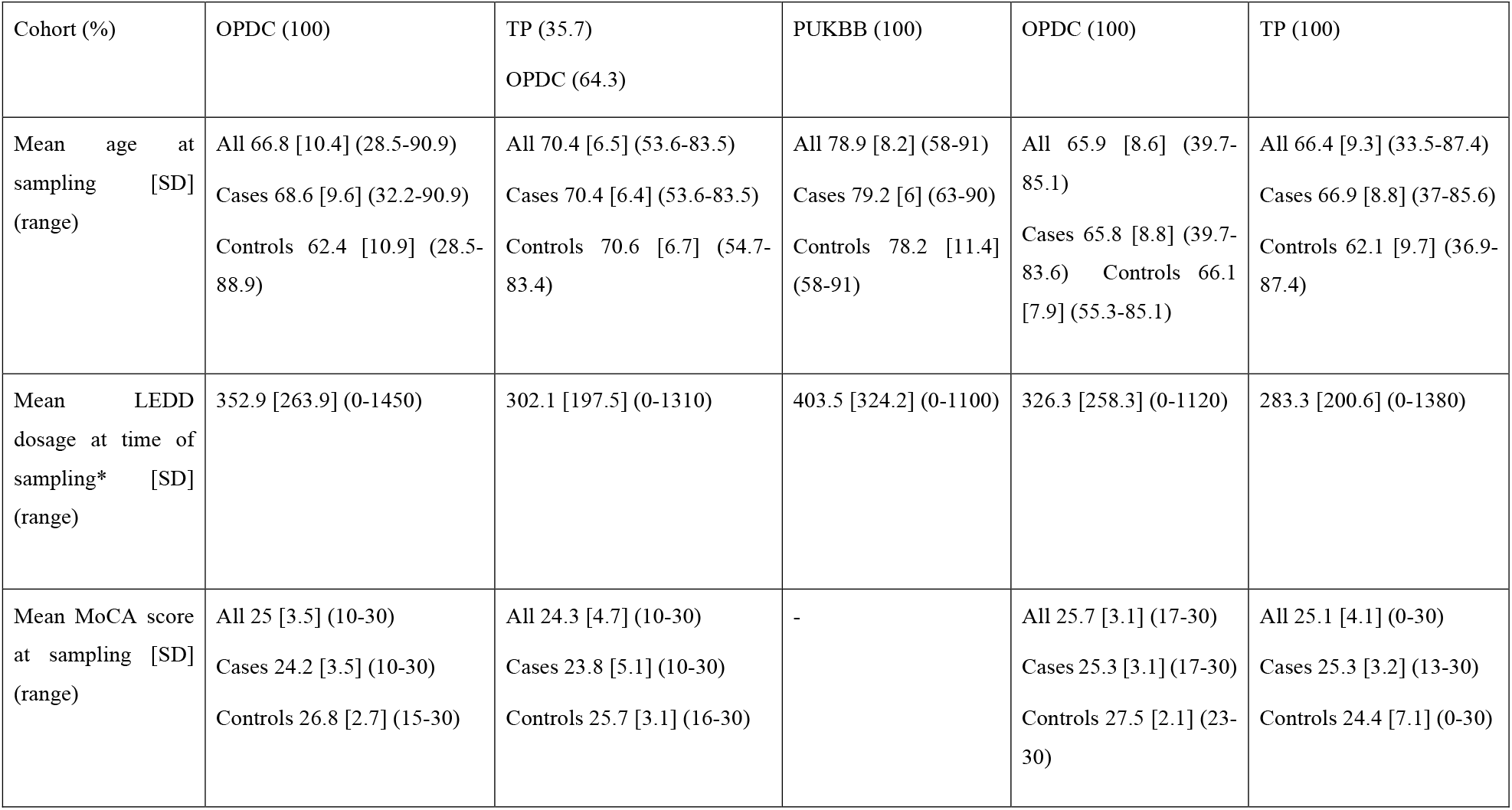
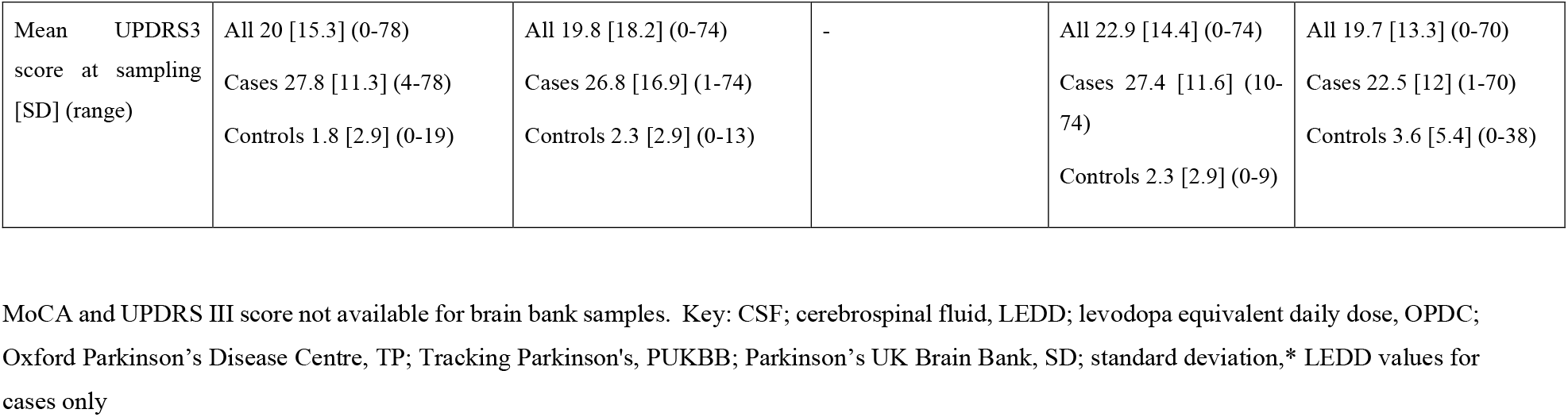
Demographics for a three cohort study of proteomic expression.

|  | OPDC - Serum | OPDC / TP - 1K | PUKBB - Brain | OPDC - CSF | TP - Serum |
| --- | --- | --- | --- | --- | --- |
| Total samples | 572 | 224 | 37 | 85 | 932 |
| Disease Status Case (%) | 400 (69.9) | 159 (71) | 24 (64.9) | 70 (82.4) | 671 (81.9) |
| Control | 172 (30.1) | 65 (29) | 13 (35.1) | 15 (17.6) | 148 (18.1) |
| Gender (%) |  |  |  |  |  |
| Male | 326 (57) | 161 (71.9) | 29 (78.4) | 53 (62.4) | 567 (60.8) |
| Female | 246 (43) | 63 (28.1) | 8 (21.6) | 32 (37.6) | 365 (39.2) |

| Cohort (%) | OPDC (100) | TP (35.7)<br>OPDC (64.3) | PUKBB (100) | OPDC (100) | TP (100) |
| --- | --- | --- | --- | --- | --- |
| Mean age at sampling [SD] (range) | All 66.8 [10.4] (28.5-90.9)<br>Cases 68.6 [9.6] (32.2-90.9)<br>Controls 62.4 [10.9] (28.5-88.9) | All 70.4 [6.5] (53.6-83.5)<br>Cases 70.4 [6.4] (53.6-83.5)<br>Controls 70.6 [6.7] (54.7-83.4) | All 78.9 [8.2] (58-91)<br>Cases 79.2 [6] (63-90)<br>Controls 78.2 [11.4] (58-91) | All 65.9 [8.6] (39.7-85.1)<br>Cases 65.8 [8.8] (39.7-83.6)<br>Controls 66.1 [7.9] (55.3-85.1) | All 66.4 [9.3] (33.5-87.4)<br>Cases 66.9 [8.8] (37-85.6)<br>Controls 62.1 [9.7] (36.9-87.4) |
| Mean LEDD dosage at time of sampling* [SD] (range) | 352.9 [263.9] (0-1450) | 302.1 [197.5] (0-1310) | 403.5 [324.2] (0-1100) | 326.3 [258.3] (0-1120) | 283.3 [200.6] (0-1380) |
| Mean MoCA score at sampling [SD] (range) | All 25 [3.5] (10-30)<br>Cases 24.2 [3.5] (10-30)<br>Controls 26.8 [2.7] (15-30) | All 24.3 [4.7] (10-30)<br>Cases 23.8 [5.1] (10-30)<br>Controls 25.7 [3.1] (16-30) | - | All 25.7 [3.1] (17-30)<br>Cases 25.3 [3.1] (17-30)<br>Controls 27.5 [2.1] (23-30) | All 25.1 [4.1] (0-30)<br>Cases 25.3 [3.2] (13-30)<br>Controls 24.4 [7.1] (0-30) |
| Mean UPDRS3 score at sampling [SD] (range) | All 20 [15.3] (0-78)<br>Cases 27.8 [11.3] (4-78)<br>Controls 1.8 [2.9] (0-19) | All 19.8 [18.2] (0-74)<br>Cases 26.8 [16.9] (1-74)<br>Controls 2.3 [2.9] (0-13) | - | All 22.9 [14.4] (0-74)<br>Cases 27.4 [11.6] (10-74)<br>Controls 2.3 [2.9] (0-9) | All 19.7 [13.3] (0-70)<br>Cases 22.5 [12] (1-70)<br>Controls 3.6 [5.4] (0-38) |

MoCA and UPDRS III score not available for brain bank samples. Key: CSF; cerebrospinal fluid, LEDD; levodopa equivalent daily dose, OPDC; Oxford Parkinson's Disease Centre, TP; Tracking Parkinson's, PUKBB; Parkinson's UK Brain Bank, SD; standard deviation,\* LEDD values for cases only

### Comparing biomarker predictions in different sample modalities

We first wanted to assess whether putative biomarkers, previously suggested from a range of published studies, could be replicated using our proteomic data. From a list of 50 PD specific, neurodegenerative and cell type markers, 25 were included in assays on the Somalogic panel. Apolipoprotein A1 (*APOA1*) and Growth Hormone Receptor (*GHR*) protein expression were related to at least one disease phenotype in all the serum sample sets tested (Figure 2). Furthermore, Bone sialoprotein (*IBSP*) was associated to the MoCA phenotype in all the serum datasets. In the CSF sample set five suggestion biomarker proteins showed an association with disease status (p-value > 0.05). These included Glial fibrillary acidic protein (*GFAP*), Bone sialoprotein (*IBSP*) and DJ-1 (*PARK7*). None of the proteins tested in brain tissue were related to disease. Interestingly, no proteins tested were significantly related to the Parkinson’s disease phenotypes in every modality. These results suggest that there is scope for improvement of single protein biomarkers, perhaps instead as part of a multi-protein panel. Therefore, the next objective was to discover new multi-protein combinations which could be utilised as a biomarker.

**Figure 2.**
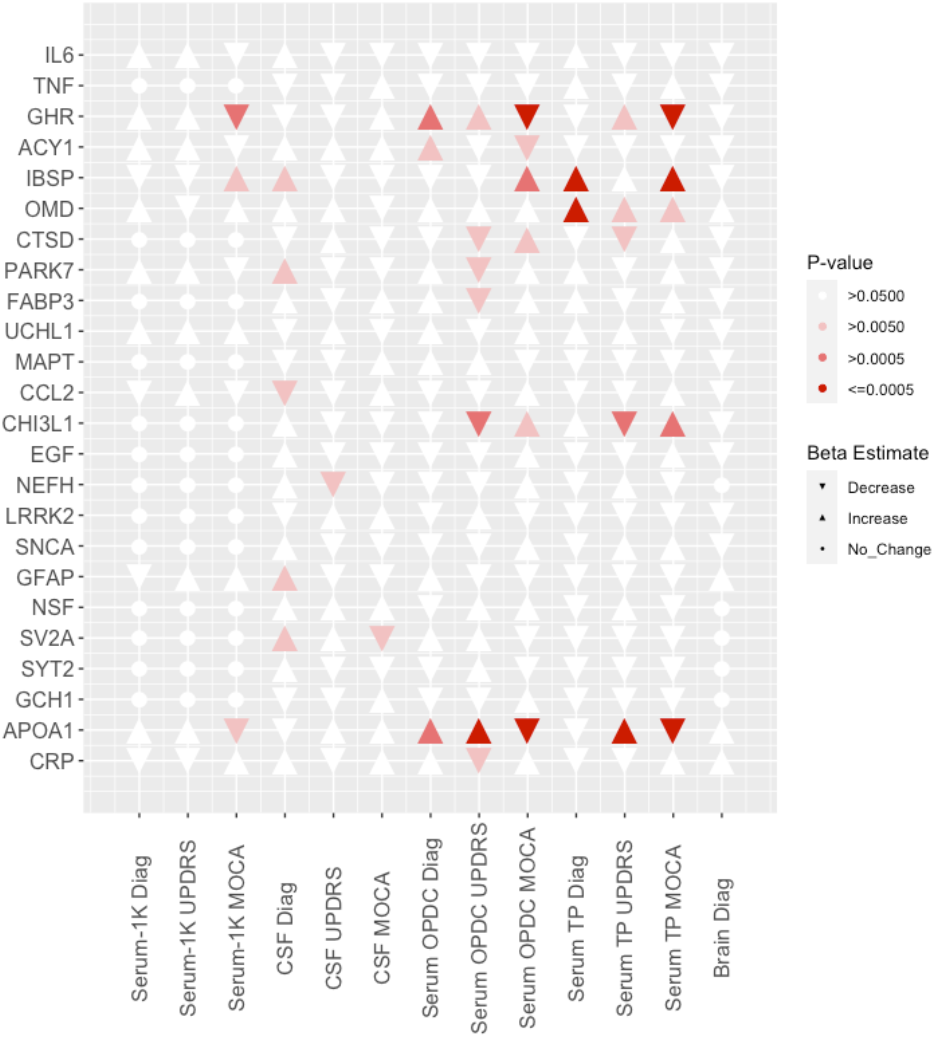
Targeted biomarker Protein Results. Significance matrix representing the association between protein expression of known biomarkers and PD phenotypes across the project datasets. Univariant logistic regression was used for diagnosis (disease status). Univariant linear regression was used for MoCA (cognition) and MDS - UPDRS (movement) scores. Direction of effect is shown by up (positive) and down (negative) triangles. Circles are used to show missing Somalogic assays.

### Defining a robust proteomic biomarker using multi-modal cohorts

Given the limited replication of previously identified biomarkers, which suggested that a known biomarker signal is detectable but perhaps not fully transferable between modalities, we went on to generate a *de novo* signature of disease from all 1,004 proteins captured by the SomaLogic data common to all three sample sources in this study. We applied machine learning approaches to first prioritise protein selection from a training set in serum (n = 572) and then carry out testing in serum (n = 227), CSF (n = 45) and brain (n = 37) samples. Briefly, we used simulated annealing and random forest for protein selection in the training serum set (OPDC - Serum) which produced 200 potential classifiers, each using 1-200 of the top-ranked proteins. In the next stage, we tested each classifier in all three modalities to understand their performance in other tissue types: post-mortem brain (PUKBB - Brain), CSF tissue (OPDC - CSF) and serum (OPDC / TP - 1K). Figure 3A shows a comparison between classifiers generated on different modality test sets. The brain sample proteins had an AUC of 0.75 (p-value=0.006), CSF AUC was 0.74 (p-value=0.0009) and serum AUC was 0.66 (p-value=0.0002). These classifiers used the top 31, 86 and 142 ranked proteins respectively (Table 2). Despite selecting from the same 200 proteins the serum sample set was optimal at 31 proteins. The best classifier outcome, optimised in the CSF samples, included APOE and GFAP (Supplementary Table 3). Enrichment analysis of the CSF classifier proteins detected four KEGG pathways including cytokine-cytokine receptor interaction (adjusted p-value=0.004) as well as complement and coagulation cascades (adjusted p-value=0.041). Using the smaller brain classifier protein list only a single KEGG pathway was found; Cytokine-cytokine receptor interaction (adjusted p-value=0.0002).

**Figure 3.**
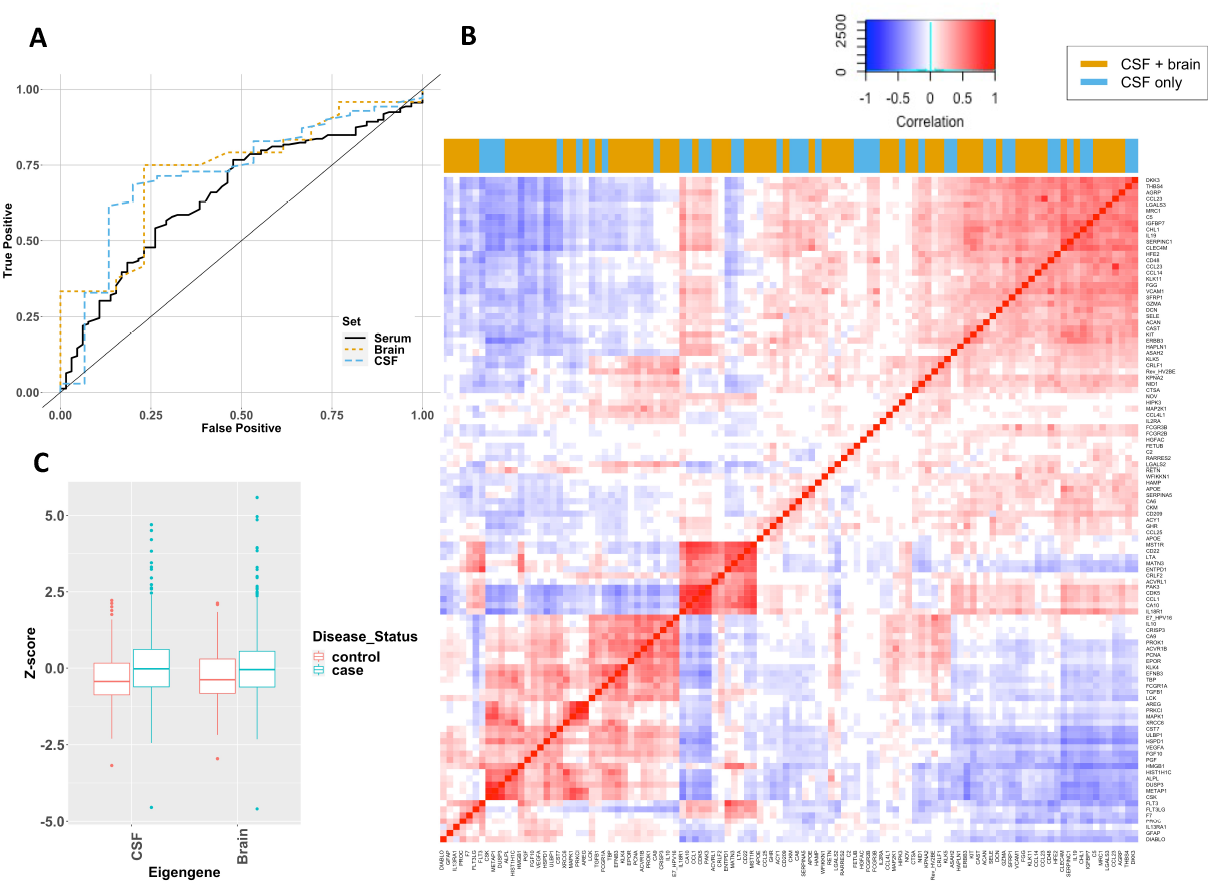
Proteins classifiers testing showed strong potential biomarkers in both CSF and brain sample sets. **A**. Receiver operator characteristic (ROC) curve for each of the best classifiers when testing in the sample sets. The black line defines what to expect by chance from a classifier. From left to right, an ideal classifier would follow a line up the y-axis then along the x-axis, which would have an AUC of 1. The testing sets are ‘Brain’, ‘CSF’ and ‘Serum’ **B**. Exploration of the signature protein expression was performed in independent TP - Serum set. Correlation between expression of the CSF classifier proteins is shown by heatmap where proteins with similar profiles are clustered together. Proteins present in both brain and CSF signatures are coloured yellow. **C**. Derived classifier Eigengenes are significantly associated to disease status in an independent serum dataset

**Table 2.** Filtering Results of top ranked proteins using three modalities. Testing classifier on three modalities to select best predictive proteins

| **Dataset** | **Number of Proteins (classifiers)** | **AUC [95% CI]** | **p-value** |
| --- | --- | --- | --- |
| **Serum** | 31 | 0.66 [0.59-0.74] | $2.00 \times 10^{-04}$ |
| **Brain** | 86 | 0.75 [0.57-0.93] | 0.0063 |
| **CSF** | 142 | 0.74 [0.61-0.87] | $9.00 \times 10^{-04}$ |

Testing classifier on different disease outcomes to identify proteins predictive of disease severity
| Dataset | Number of Proteins (classifiers) | AUC [95% CI] | p-value |
| --- | --- | --- | --- |
| Severe | 31 | 0.72 [0.63-0.81] | 1.0x10<sup>-04</sup> |
| Intermediate | 18 | 0.72 [0.61-0.83] | 2.0x10<sup>-04</sup> |
| Mild | 12 | 0.63 [0.53-0.72] | 0.0063 |

### Validation of the novel protein panel in an independent cohort

Finally, we sought to validate the best of the classifiers using a set of serum samples (n=932) from the Tracking Parkinson’s cohort that had not been included in the prior training and testing process. Given the similarity between AUC (0.74 and 0.75) of the CSF and brain classifiers we took both forward for further analysis. Initially, we used exploratory methods to understand how the classifier proteins related to each other. Focusing first on the larger protein signature, protein expression was tested for between protein correlations to understand the range and representation. Figure 3B shows the clustering of the correlation values to illustrate the similarities between subgroups of proteins in the signature. In order to validate the complex signature of disease, we converted the classifiers to a single indicator or eigengene. This single value expression signature was calculated from a mean per sample as well as a dimensional reduction approach to produce an eigengene per sample. These single value expression signatures were compared to clinical outcomes (Figure 3C). Both the CSF and brain classifiers showed higher values and were significantly associated with diagnosis status using both signature methods (p-values < 0.001, Supplementary Table 4) providing further validation.

### Understanding the effects of disease severity on biomarker discovery

To understand the effects of disease severity we then compared the effects on the protein signature on a defined subgroup of patients with relatively severe and mild disease phenotypes. We tested our 200 protein classifier (as defined previously) in three serum sample subsets (severe, intermediate and mild). These independent serum classifiers demonstrated good predictive performance in finding the difference between outcome sample set as compared to the control group (p values < 0.01 Table 2B). The severe outcome subgroup had an AUC of 0.72 with a selection of 31 proteins. This is improved compared to the AUC for the complete serum disease cohort of 0.66. These 31 proteins were enriched only for the Cytokine-cytokine receptor interaction; 12 of the proteins being in this KEGG pathway (adjusted p-value=0.0018).

As the strongest classifier, the severe outcome signature was explored further in the independent serum validation set (Tracking Parkinson’s). We compared the 22 proteins passing cohort-level quality control to the PD phenotypes to understand whether the severe outcome proteins reflected disease severity in an alternative dataset. The MDS-UPDRS III and severity scores were significantly associated to the severe phenotype protein signature (beta = -0.13, p-value=0.006 and beta=-0.14, p-value=0.025). There was no significant association between MoCA and the severe phenotype signature.

### Differential expression of key proteins demonstrates a relationship to both known and novel PD mechanisms

Having identified a possible biomarker signature from the 1,004 proteins common across the samples we used, we then went on to explore protein differences using the full 4,001 proteins in samples analysed with version 4 of the SomaLogic panel. A total of 1,599 serum samples were analysed with the Version 4 array from which 3,378 proteins passed QC in all samples.

We performed univariable regression to ascertain the significance of each protein individually. This was implemented for each of the outcome variables; disease status (case vs. control), cognitive decline (MoCA score) and motor symptoms (MDS UPDRS III score). The greatest changes in protein expression were found when comparing the disease status of the samples and this resulted in 261 differentially expressed proteins used for pathway analysis (Figure 4A). In addition, 89 proteins were associated with motor function (MDS-UPDRS III) and 13 with cognition (MoCA). Three proteins were associated with all phenotypes: Oncostatin-M (*OSM*); Complement component 9 (*C9*) and Carnosine Dipeptidase 1 (*CNDP1*).

**Figure 4.**
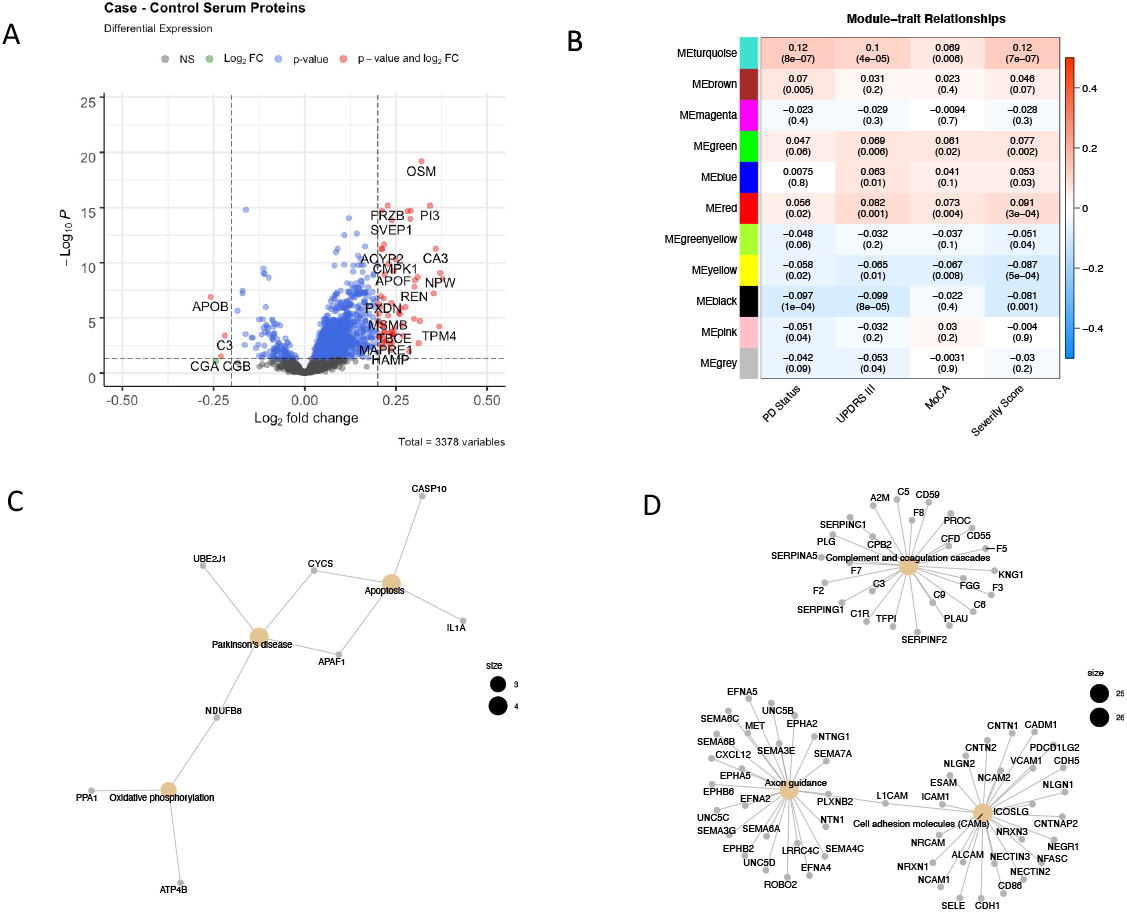
Differential Regulation and WGCNA summary from Serum Protein Expression. **A**. Fold Change Plot showing genes with differential regulation between PD case vs control **B**. WGCNA Module Eigengenes correlation comparison to PD traits **C**. Yellow Module Protein Network with known PD protein pathway focus **D**. Turquoise Protein Network includes pathways with immune function and axon guidance

To understand whether the changes in protein could potentially show changes in function we performed enrichment analysis. Pathway analysis showed that the main significantly dysregulated protein pathways of case status were the complement and coagulation cascades (p-value = 0.009) and Protein digestion and absorption (p-value = 0.015). The Fat digestion and protein absorption pathway was enriched for dysregulated proteins associated with motor symptoms as measured using the MDS-UPDRS III (p-value = 0.001).

We then used co-expression analysis with WGCNA to identify modules of differential protein expression associated with disease status within each case a protein eigengene to represent that module expression. Four module eigengenes were significantly correlated to all PD phenotypes (Figure 4B p-value < 0.05). The yellow module eigengene was negatively correlated to the tested traits. It contained only 98 proteins and had a relationship directly to disease pathways (PD, Oxidative phosphorylation and apoptosis, p-value < 0.05, Figure 4C). The largest module (turquoise) was positively correlated to the traits and had 704 proteins and three key disease pathways including complement and coagulation cascades and axon guidance (Figure 4D). Interestingly, the red module was enriched for immune pathways in both the GO and KEGG databases (Supplementary Table 2). The green module had little apparent functional relevance. Table 3 shows details for the three significant disease related modules. Each module has a hub protein which has the strongest association with the rest of the module. The turquoise module is represented by the Neurogenic locus notch homolog protein 1 precursor (*NOTCH1*) protein, part of the Notch signalling pathway which is involved in vascular development and angiogenesis. 15-hydroxyprostaglandin dehydrogenase (*HPDG*), the hub protein for the red module is part of the prostaglandin signalling pathway.

**Table 3.**
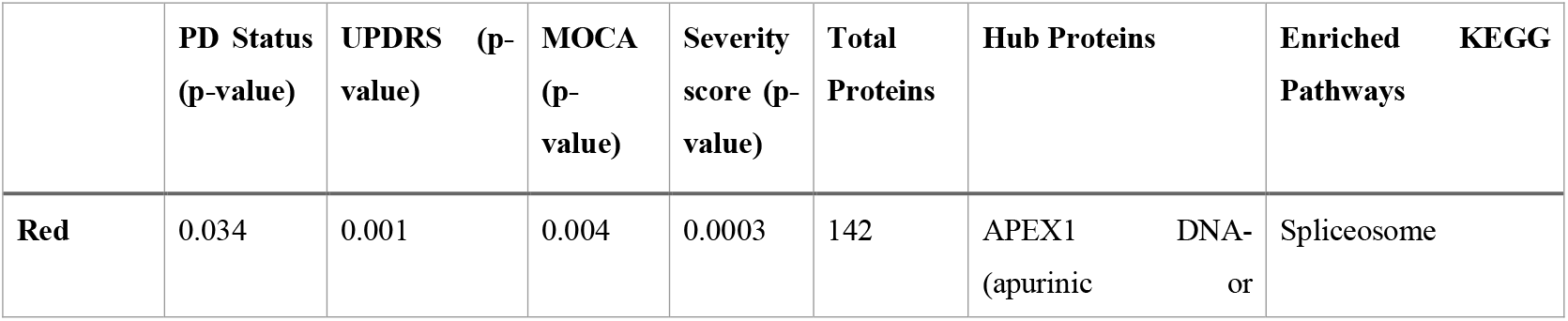
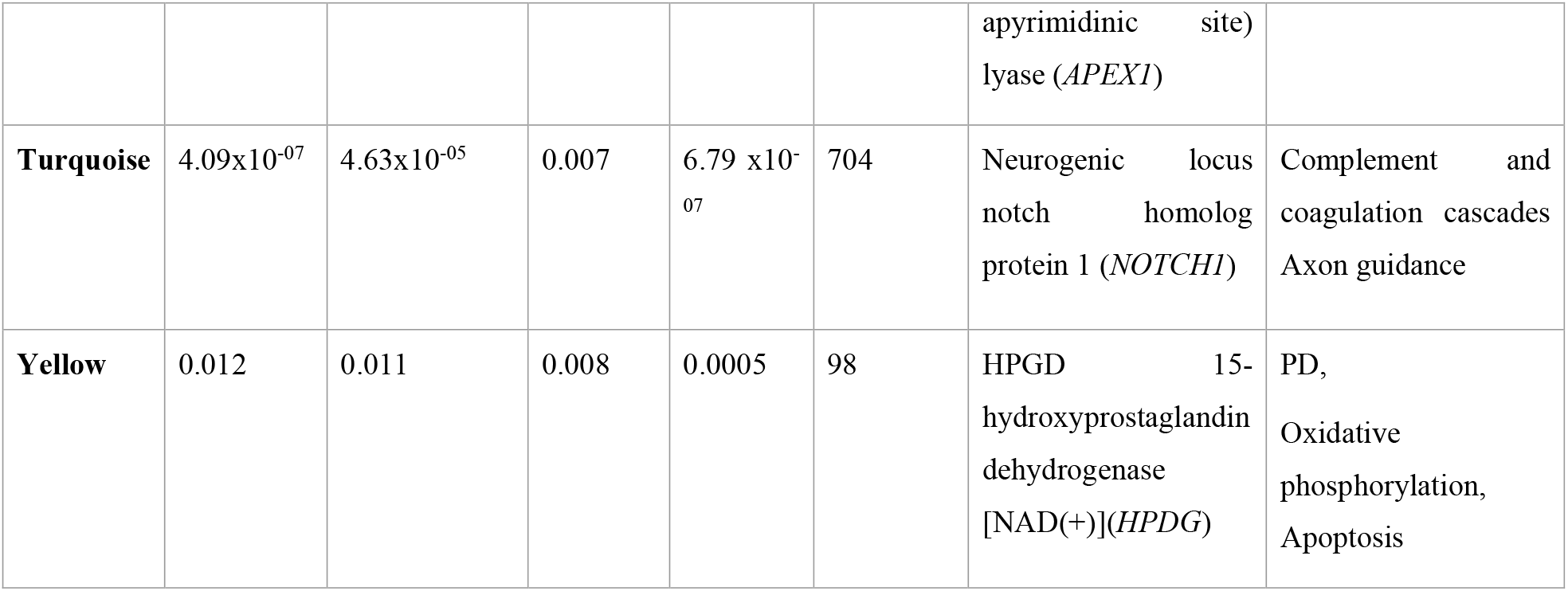
WGCNA Protein Modules with endophenotype relationships. Modules with significant correlation with all disease phenotype were selected for further analysis. The three modules described passed p-value thresholds (adjusted p-value < 0.05) and had functional relevance.

## Discussion

### Key Results

We report here a large biomarker discovery programme including both unsupervised and confirmatory analyses using three different modalities (serum, CSF and brain tissue) and three different cohorts with a panel of aptamer capture protein assays including over 1,000 and 4,000 proteins in different phases of the study.

The first analysis generated a novel signature of proteins associated with PD. We used a machine learning approach to identify a protein signature differentiating case samples from control subjects. The resulting classifier was tested on two different cohorts to produce a more robust signature. Testing was also performed across tissue modalities to produce a more representative signature of disease (AUC = 0.66 to 0.75). Reproducibility was a priority for study design hence the inclusion of a comprehensive serum validation set where the protein signature had a significant association to PD status and movement phenotypes (p-values < 0.001). This final signature contained proteins from the cytokine-cytokine receptor interaction pathway as well as the complement pathway. Differential regulation and clustering of protein expression provided further evidence for enrichment for the complement pathway in PD; a key element of the innate immune response that appears to be a common feature of multiple neurodegenerative diseases [24]. Findings are consistent with previous work from the OPDC cohort showing that the most severely affected baseline PD subtype was associated with a proinflammatory profile, comprising raised *CRP* and reduced *APOEA1* [25].

The AUC values reported here are modest and lower than would be hoped for a simple blood-based biomarker to be used in clinical practice, albeit that the CSF values were reasonable. However, the fact that we show robust replication and validation across multiple tissue types and cohort sources demonstrates that a blood-based signature of PD is an achievable objective. Moreover, the fact that we identify pathways and processes known to be involved in neurodegeneration including PD strongly supports the contention that this signature is biologically relevant. Further work will be needed to identify the optimal components of this signature for use in practice in clinical research and technological enhancements of future arrays may have better predictive value.

A biomarker would have the most clinical utility if it could be used for early, ideally preclinical, identification of disease or if it could be used as a surrogate for more specific markers of disease pathology. In the related field of AD biomarkers, the availability of definitive markers of disease pathology such as PET and CSF markers of both amyloid and tau pathology have enabled such studies. For example, PET Amyloid can be used as an endpoint or outcome variable for biomarker discovery [13,26]. However, such designs are not yet possible in PD – there is no PET radioligand or CSF assay of alpha-synuclein to act as the equivalent anchor point. In the absence of such in vivo markers of disease pathology, we used a design where we compared different stages of disease using measures of motor and cognitive progression in the hope that a change in biomarker across people with symptomatic disease could be extrapolated to very early or prodromal disease and potentially to truly preclinical disease. Using this approach to describe the phenotypes and building on the ML methods we generated a serum protein signature tested in severe, mild and intermediate patient subsets. The classifiers had an increased AUC (0.72) compared to the ungrouped serum samples (0.66) suggesting a stricter disease phenotype is better for disease prediction and therefore refining the proteins included in the signature.

The signature for ‘severe’ disease included particular proteins related to cytokine receptor enrichment which constituted nearly half of the signature. These proteins were associated with both severity and motor phenotypes in the validation analysis. Although we do not have a clear biomarker for preclinical changes we have been able to use the approach to understand more about the involvement of key proteins in disease progression. Further studies to examine this signature, and its constituent proteins, in early disease and in preclinical and prodromal PD cases from longitudinal studies are warranted and will confirm if they can be used to predict the rate of conversion to PD clinical diagnosis.

There are several novel proteins described in these analyses which are common to more than one approach. Oncostatin-1 (*OSM*) was found in the CSF classifier and also detected in the analyses of differential regulation in relation to disease status, cognitive and movement scores. The known biomarkers APO1A [27] and GFAP were present in the CSF classifier which could act as positive controls for the signature. Using pathway enrichment approaches we identified many PD relevant pathways. The axon guidance pathway contains proteins related to synaptic dysfunction [28] and the complement and coagulation cascades are an important pathway in innate immune responses previously identified in neurodegenerative diseases [24,29]. Using co-expression clustering we detected a module (turquoise) represented by the NOTCH1 protein, which has a reported PD relationship as it is modulated by the LRRK2 protein [30]. HPDG, the hub protein for the red module is part of the prostaglandin signalling pathway with a role in neurodegeneration.

### Limitations

As in any observational cohort study, there are limitations of this work. We used two cohorts, one brain bank and three sample modalities, which although gave great power to detect associations also introduced cohort and tissue noise. Without pathological confirmation, there is likely to be some misclassification due to cases of multiple system atrophy or progressive supranuclear palsy being wrongly included. This is likely to be a low rate as clinicians were all movement disorder specialists [31], red flags were used to exclude participants, we excluded those with a probability of PD < 90% and follow-up allowed us to check for revised patient diagnoses. Additionally, the structure of the TP cohort is different to the OPDC cohort. The relationship between case and controls in TP, although randomised, removes the traditional assumption of independence between groups. However, the strength of validating a signature in a differently structured dataset should not be overlooked and this gives a more robust final result.

### Interpretation

Here, we report a proteomic signature of PD in the blood that replicates across cohorts and validates across tissue types. Although of modest predictive power, the significance and replicability of the result is encouraging in the search for biomarkers that could be used to enhance clinical research, specifically trials, to identify and recruit well-characterised participants to clinical trials. Deep proteomic analyses of over 4,000 proteins identified disease pathways, including those of neuroimmunity and other known neurodegeneration associated functions, suggesting that targeting some of these biological processes might be of value in disease modification therapeutics. Indeed, these results raise the possibility of selecting subgroups of people for clinical trials using a precision medicine approach. Interestingly, the overlap in disease process between PD associated proteins in this study and AD associated proteins in similar studies in that disease again points to commonalities and overlaps between neurodegeneration suggesting that a precision medicine approach might not be possible but might be a necessary route to effective treatments for these devastating disorders.

## Conclusions

Using the largest and most comprehensive proteomics PD datasets, we have described a range of approaches for biomarker discovery and confirmed the potential of large-scale data analysis. We have demonstrated the potential value of a multi-protein signature of novel markers for further analysis and replication in new large datasets as these become available.

## Data Availability

Data is available on application to cohort owners.

## Additional Information

## Acronyms

AD: Alzheimer’s Disease
CSF: Cerebrospinal fluid
ML: Machine learning
MoCA: Montreal Cognitive Assessment
OPDC: Oxford Parkinson’s Disease Centre
PD: Parkinson’s Disease
PUKBB: Parkinson’s United Kingdom Brain Bank
TP: Tracking Parkinson’s
WGCNA: Weighted Gene Correlation Network Analysis
MDS - UPDRS: Movement Disorder Society - Unified Parkinson’s Disease Rating Scale

## Acknowledgements

We would like to thank the OPDC, PUKBB and Tracking Parkinson’s cohort participants.

## Author contributions

Original idea was conceived by SL. Study plans were developed by SL, MH, YBS, RWM, DG, JA, IM, and LW. Data analysis was carried out by LW, ML, IM, JA, BL, ANH. LW, IM and SL drafted the initial manuscript with all other authors. All authors reviewed the final manuscript.

## Ethics / Patient consent

The Oxford Parkinson’s Disease Centre study was undertaken with the understanding and written consent of each subject, with the approval of the National Research Ethics Service (NRES) Committee South Central – Oxford A (Ref:16/SC/0108); Oxford C (Ref:15/SC/0117) and Berkshire Research Ethics Committee (Ref: 10/H0505/71). This was in compliance with national legislation and the Declaration of Helsinki.

The Tracking Parkinson’s study is carried out in accordance with the Declaration of Helsinki. Patient consent was obtained and ethics approval was made by the West of Scotland Ethics Committee.

The Parkinson’s UK Brain Bank is part of the Multiple Sclerosis Society and Parkinson’s Tissue Bank at Imperial College London. It has been approved as a research tissue bank by the Wales Research Ethics Committee (reference number 08/mre09/31+5).

## Data availability

Data is available on application to cohort owners.

